# Analysis of fragment ends in plasma DNA from patients with cancer

**DOI:** 10.1101/2021.04.23.21255935

**Authors:** Karan K. Budhraja, Bradon R. McDonald, Michelle D. Stephens, Tania Contente-Cuomo, Havell Markus, Maria Farooq, Patricia F. Favaro, Sydney Connor, Sara A. Byron, Jan B. Egan, Brenda Ernst, Timothy K. McDaniel, Aleksandar Sekulic, Nhan L. Tran, Michael D. Prados, Mitesh J. Borad, Michael E. Berens, Barbara A. Pockaj, Patricia M. LoRusso, Alan Bryce, Jeffrey M. Trent, Muhammed Murtaza

## Abstract

Fragmentation patterns observed in plasma DNA reflect chromatin accessibility in contributing cells. Since DNA shed from cancer cells and blood cells may differ in fragmentation patterns, we investigated whether analysis of genomic positioning and nucleotide sequence at fragment ends can reveal the presence of tumor DNA in blood and aid cancer diagnostics. We analyzed whole genome sequencing data from >2700 plasma DNA samples including healthy individuals and patients with 11 different cancer types. We observed higher fractions of fragments with aberrantly positioned ends in patients with cancer, driven by contribution of tumor DNA into plasma. Genomewide analysis of fragment ends using machine learning showed overall area under the receiver operative characteristic curve of 0.96 for detection of cancer. Our findings remained robust with as few as 1 million fragments analyzed per sample, suggesting that analysis of fragment ends can become a cost-effective and accessible approach for cancer detection and monitoring.

**One-sentence summary:** Analyzing the positioning and nucleotide sequence at fragment ends in plasma DNA may enable cancer diagnostics.

---

Analysis of plasma DNA has enabled novel diagnostic approaches in prenatal(*1*), transplant(*2*) and cancer medicine(*3*). Recent studies have shown fragmentation patterns in cell-free DNA are not random and reflect chromatin accessibility in the cells that contribute such DNA into plasma(*4*). DNA fragments from genomic loci bound by nucleosomes or other proteins are protected from degradation in plasma(*5*). Nucleosome positioning and chromatin accessibility across the genome vary between cell types and in different cell states(*6*). Reflecting this variation, when DNA from a cancer cell is shed into plasma, the protected fragments may differ in genomic position relative to the majority of cell-free DNA in plasma which is derived from peripheral blood cells(*7*). Here, we evaluate the hypothesis that differences in fragmentation breakpoints of tumor-derived DNA in plasma can serve as a cancer biomarker, using genomewide analysis of positioning and nucleotide sequence at fragment ends in plasma DNA.

To evaluate whether genomic positioning of fragment ends in plasma DNA was different between cancer patients and healthy individuals, we first inferred a map of genomic regions recurrently protected from degradation using whole genome sequencing of plasma DNA from 17 healthy individuals. We then quantified the fraction of fragments whose ends fall within these recurrently protected regions in plasma samples from cancer patients and healthy individuals (Fig. 1A and table S1). Compared to 40 plasma samples from healthy individuals, the mean fraction of these aberrant fragments (FAF) was higher in 261 samples from patients with melanoma (P = 3.3 × 10^−6^), 46 samples from patients with cholangiocarcinoma (P = 9.4 × 10^−11^), 45 samples from patients with glioblastoma (P = 9.3 × 10^−9^) and 47 samples from patients with breast cancer (P = 4.5 × 10^−8^). To determine whether FAF was related to fraction of tumor DNA in plasma, we compared FAF with tumor fraction measured using analysis of copy number aberrations in patients with advanced cancer. FAF was correlated with tumor fraction in patients with melanoma (r = 0.76, P = 9.9 × 10^−51^; Fig. 1B), and in patients with cholangiocarcinoma (r = 0.71, P = 2.2 × 10^−8^; fig. S1). In longitudinal samples from patients with metastatic melanoma, changes in FAF during therapy were consistent with changes in tumor fraction in plasma DNA (Fig. 1C and fig. S2). In some cases, changes in FAF were concordant with treatment response on imaging, even when tumor fraction in plasma DNA was undetectable using copy number analysis. In 3 patients with glioblastoma, we compared FAF with tumor fraction in plasma DNA measured using targeted digital sequencing(*8*). We found that changes in FAF during therapy were consistent with changes in tumor fraction, even though detectable circulating tumor DNA concentrations were very low (0.01% to 1.2%; Fig. 1D and fig. S3). To ascertain whether aberrant DNA fragments in plasma were contributed by the tumor, we focused our analysis on patients with metastatic melanoma and samples with high tumor fraction in plasma DNA. Since the tumor contributes more fragments of plasma DNA from genomic loci that are gained in the tumor genome compared to those that are lost, we expected FAF to be higher for genomic loci affected by copy number gains if tumor-derived DNA fragments in plasma are aberrant. In 27 plasma samples with at least 20% tumor fraction in plasma, we found that FAF was higher at genomic loci affected by copy number gains compared to loci unaffected by copy number changes or those affected by copy number losses (Fig. 1E and fig. S4). To further assess the tumor specificity of aberrant DNA fragments in plasma, we performed deep whole genome sequencing at >280x coverage for two plasma samples with tumor fractions of 36% and 39%. We evaluated whether DNA fragments carrying tumor-specific mutations were more likely to be aberrant (Fig. 1F and table S2). In both plasma samples, we found that a greater fraction of mutated fragments were aberrant compared to non-mutated fragments covering the same genomic loci (P < 2.2 × 10^−16^ and P = 1.5 × 10^−11^). Taken together, these results demonstrate that elevated fraction of aberrant fragments observed in plasma DNA from patients with cancer is driven by tumor-derived DNA fragments.

**Fig. 1.**
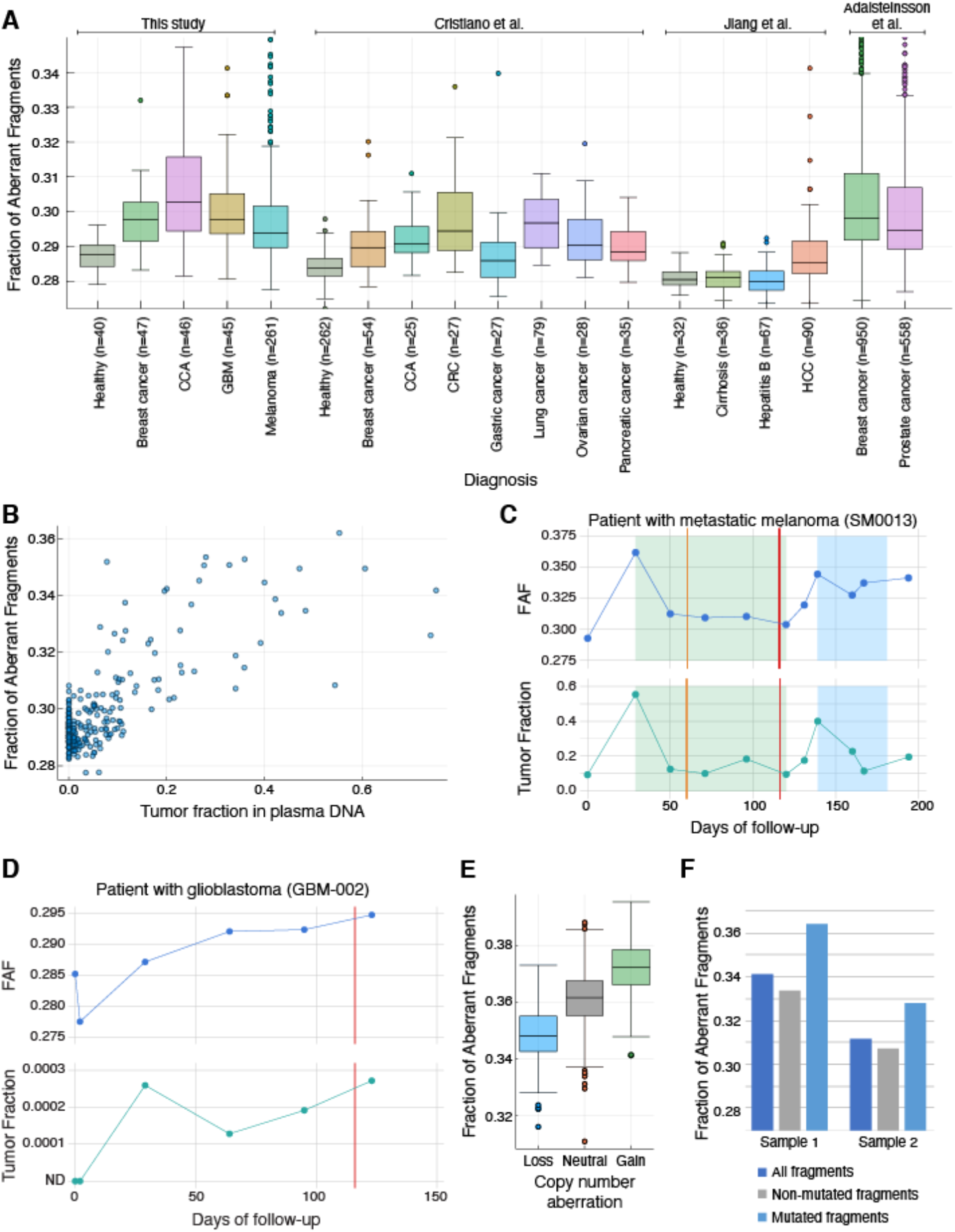
Fraction of aberrant fragments in plasma samples from patients with cancer. FAF was higher in plasma samples from patients with cancer compared to healthy volunteers, in whole genome sequence data from >2700 plasma samples (**A**). FAF was correlated with tumor fraction measured using copy number analysis in plasma samples. Results from patients with metastatic melanoma are shown in (**B**) and additional results are shown from patients with cholangiocarcinoma (fig. S1), breast cancer and prostate cancer (fig. S5). Longitudinal changes in FAF during therapy were consistent with changes in tumor fraction measured by copy number analysis in patients with metastatic melanoma. Results from a representative patient are shown in (**C**). Upper panel shows changes in FAF over time and lower panel shows changes in tumor fraction measured using copy number analysis by ichorCNA. Shaded regions of the plot show on-going systemic therapy. Orange vertical line indicates Stable Disease on imaging and red vertical line indicates Progressive Disease on imaging. Results from additional patients are shown in fig. S2. Despite very low tumor fractions observed in patients with glioblastoma, longitudinal changes in FAF during therapy were consistent with changes in tumor fraction measured using targeted digital sequencing. Results from a representative patient are shown in (**D**) and results from additional patients are shown in fig. S3. The first sample was collected prior to surgical resection of the tumor and subsequent samples were collected after surgery during therapy. Vertical red line indicates clinical progression of disease. FAF was higher at genomic loci affected by copy number gain in the corresponding tumor genome, compared to unaffected loci or those affected by copy number loss. Results from a representative patient with metastatic melanoma are shown in (**E**) and results from additional patients are shown in fig. S4. For two plasma samples with higher tumor fraction in plasma, we compared FAF between mutated and non-mutated fragments and these results are shown in (**F**). FAF for mutated DNA fragments was higher than non-mutated DNA fragments (Table S3). CCA, cholangiocarcinoma. GBM, glioblastoma. CRC, colorectal cancer. HCC, hepatocellular carcinoma.

To evaluate external validity of our findings and ensure our observations were not driven by artifacts arising from sample handling and processing, we analyzed fragment ends in whole genome sequencing data from 3 recent publications, including 2270 plasma DNA samples from patients with cancer and healthy individuals(*9-12*). We found FAF was elevated across multiple cancer types and stages (Fig. 1A and table S2). In patients with metastatic breast and prostate cancer with high circulating tumor DNA concentrations, FAF was correlated with tumor fraction in plasma DNA (fig. S5). Plasma samples from patients with liver cirrhosis or hepatitis were not distinguishable from healthy individuals. However, plasma samples from patients with liver cancer showed higher FAF compared to healthy individuals, patients with liver cirrhosis and patients with hepatitis (P < 0.001; Fig. 1A). These results showed that analysis of fragment ends can be generalized across cancer types and is relevant even for datasets that were generated independently from our laboratory.

Using our approach for calculation of FAF, whether a fragment is aberrant can only be determined if it intersects with a genomic locus known to be protected from degradation in healthy individuals. This approach excludes any fragments that map to other unannotated regions of the genome, limiting the proportion of informative data to a mean of 34% from plasma DNA samples. Differences in genomic positioning of plasma DNA fragments can also be captured through analysis of nucleotide sequence at fragment ends. To analyze fragment ends from all mapped DNA fragments, we investigated nucleotide frequencies observed at each position 10 bp upstream and downstream of each fragment end (based on the reference genome sequence), averaged across all fragments for each sample. To assess whether differences in average nucleotide frequencies at fragment ends were driven by tumor contribution in plasma DNA, we used multidimensional scaling and compared the first two dimensions of nucleotide frequencies with FAF and with tumor fraction for 4 cohorts of patients with advanced cancers and high circulating tumor DNA concentrations. Absolute values for correlation between the second dimension of nucleotide frequencies at fragment ends and FAF were 0.70, 0.73, 0.71 and 0.48 for patients with melanoma, cholangiocarcinoma, breast cancer and prostate cancer, respectively. Correlation between nucleotide frequencies at fragment ends and tumor fraction in plasma was 0.55, 0.55, 0.57 and 0.45, respectively (table S3). These results show that differences in average nucleotide frequencies at fragment ends are, at least in part, driven by tumor contribution of aberrant fragments into plasma DNA.

To evaluate whether genomewide analysis of fragment ends allows detection of cancer, we trained a machine learning model based on random forests to classify plasma samples from cancer patients and healthy individuals. The model leverages information provided by both FAF and nucleotide frequencies at fragment ends. To avoid overfitting our classification model, we restricted the analysis to the earliest available plasma sample for each patient (generally obtained at enrollment in the clinical study). This analysis was averaged over 100 runs, using 80% of samples for training and 20% for testing in each iteration (split proportionally for each cancer type and cohort). Across all patients in our cohort, analysis of fragment ends achieved an area under the receiver operating characteristic curve (AUC) value of 0.96 (Fig. 2A). Performance varied across cancer types and AUC values for melanoma, cholangiocarcinoma, breast cancer, and glioblastoma were 0.94, 0.99, 0.95, and 0.98 respectively (Fig. 2B). At 95% specificity, sensitivity for cancer detection was 79% for all cancer types, 68% for melanoma, 78% for breast cancer, 99% for glioblastoma and 99% for cholangiocarcinoma. To evaluate whether analysis of fragment ends was generalizable for cancer detection beyond our own samples, we implemented this approach using plasma whole genome sequencing data from a recent publication(*12*) that included a greater number of healthy individuals, multiple cancer types and more samples from patients with stage I-III cancers. Following a similar setup for training and testing, we found an AUC value of 0.94 across all patients (Fig. 2C). At 95% specificity, sensitivity for cancer detection was 76% for all cancer types, 59% for ovarian cancer, 67% for pancreatic cancer, 75% for breast cancer, 80% for colorectal cancer, 86% for gastric cancer, 90% for cholangiocarcinoma and 97% for lung cancer (fig. S6). When this analysis was limited to patients with potentially curable Stage I-III disease, AUC value dropped marginally to 0.93, with 75% sensitivity at 95% specificity (Fig. 2D).

**Fig. 2.**
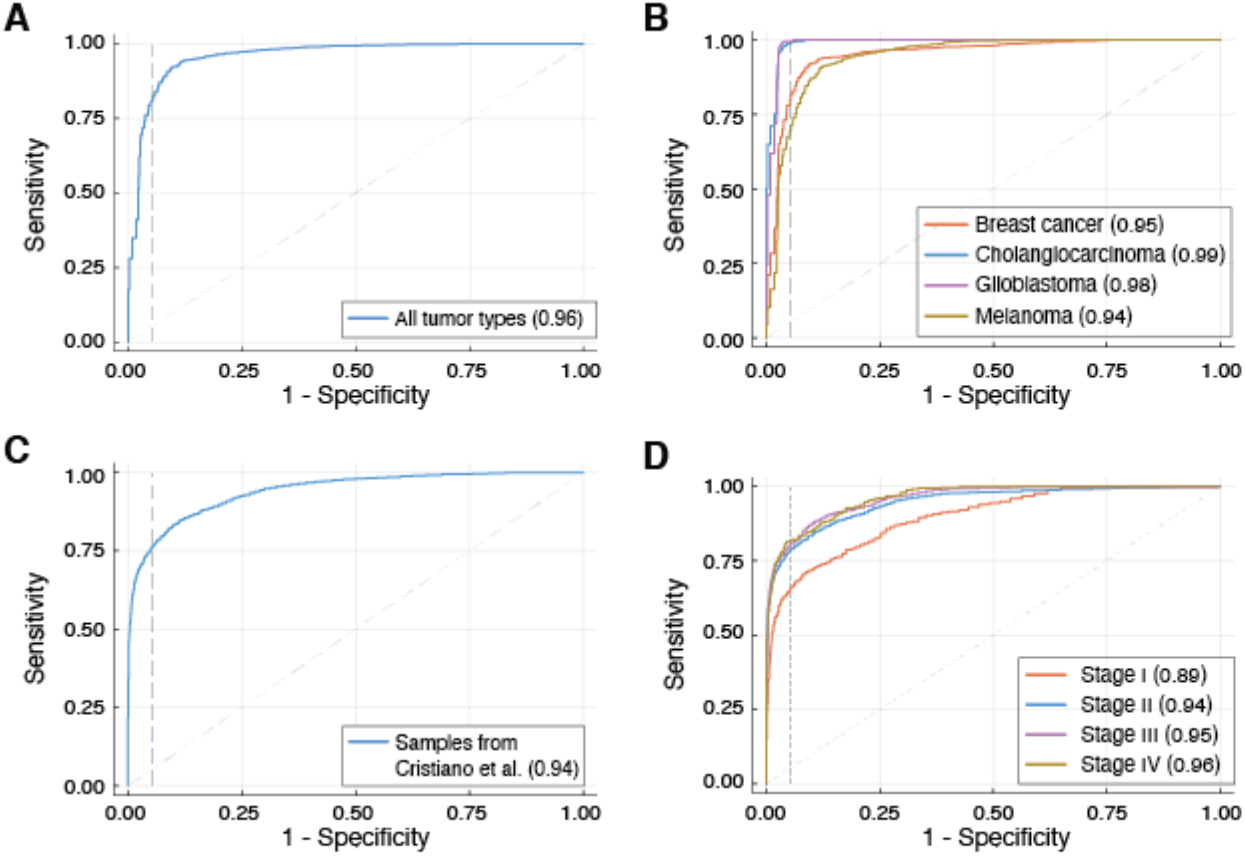
Diagnostic performance for cancer detection using analysis of fragment ends. Results from a random forests classifier trained to distinguish cancer patients from healthy individuals, using fraction of aberrant fragments and average nucleotide frequencies at fragment starts and ends in plasma whole genome sequencing data. For samples in our cohort, overall performance is shown in (**A**), and performance by tumor type is shown in (**B**). For samples in Cristiano et al. (*12*), overall performance is shown in (**C**), and performance by disease stage is shown in (**D**).

To determine whether analysis of fragment ends in plasma DNA is robust at shallow sequencing coverage, we down-sampled sequencing data, and evaluated measurement of FAF and performance of the classifier at multiple read depths. Even when only a maximum of 1 million fragments were analyzed, we found that co-efficient of variation for FAF was less than 1% in independent sets of fragments from the same plasma samples (fig. S7). When fragments per sample were limited to 1 million, AUC values obtained for the random forests classifier were 0.87 for our multi-cancer cohort and 0.93 for published data(*12*), respectively (fig. S8 and fig. S9).

Overall, our results demonstrate that across multiple cancer types, ends of tumor-derived plasma DNA fragments are more likely to be observed at different genomic loci compared to ends of background DNA fragments contributed by peripheral blood cells. We leveraged this observation and showed proof-of-principle results that analysis of fragment ends can be useful as a biomarker for cancer detection and monitoring of treatment response. This approach appears potentially useful for several cancer types where detection of cancer at earlier stages could improve outcomes but there are no established methods for screening, including cholangiocarcinoma, pancreatic cancer, gastric cancer and ovarian cancer. In addition, the diagnostic performance of this approach in patients with glioblastoma is particularly surprising, given how challenging circulating tumor DNA detection has been for these patients using mutation-based assays. A potential explanation for this finding is that analysis of fragment ends leverages differences in cell-free DNA shedding from different tissue types in healthy individuals and patients with cancer. Hence, this approach may perform better for cancers originating in tissues that rarely contribute cell-free DNA into plasma within healthy individuals. However, this also suggests a potential limitation that aberrant fragmentation patterns in plasma may not be specific to cancer and may arise from unexpected tissue contributions in plasma due to other systemic or acute conditions including pregnancy and transplant(*13*). In our analysis, we did not find elevated FAF in plasma samples from patients with liver cirrhosis or hepatitis, and a higher FAF was observed in patients with hepatocellular carcinoma. To utilize this approach for cancer detection, a reference dataset may be needed that includes healthy individuals across age, gender and co-morbidities. In addition, each patient’s results may need to be obtained when they are unaffected by acute illness and interpreted in the appropriate clinical context. Our approach can be improved further through analysis of even larger number of samples from patients across disease stages for each cancer type to increase accuracy of cancer detection. In the future, such data may also be useful to predict tumor type for plasma samples from cancer patients, either through selection of the most informative genomic regions to calculate FAF, and by identifying cancer type-specific nucleotide motifs and frequencies at fragment ends.

Earlier studies of fragmentation patterns in circulating tumor DNA have evaluated local differences in average fragment size in windows across the genome as an independent approach for cancer detection(*12*), or used fragment size to improve sensitivity for detection of somatic genomic alterations(*14, 15*). Another study identified genomic loci(*16*) and nucleotide motifs(*13*) preferentially utilized by DNA shed from liver cells and found liver-derived DNA was higher in patients with hepatocellular carcinoma. In contrast to these studies, we have found that fraction of aberrant fragments and average nucleotide frequencies at fragment ends, measured in aggregate for each sample, can serve as a biomarker for multiple cancer types. Analyzing just one million fragments per sample from plasma whole genome sequencing libraries, we found that the performance of our approach for cancer detection parallels published methods based on more complex analysis of mutations(*17*), methylation(*18*) and fragment size(*12*) that require higher amounts of input DNA or greater depth of sequencing. The simplicity of our approach as well as the small amount of plasma DNA and sequencing data required can greatly increase access to blood-based cancer detection and monitoring, particularly for resource-constrained health systems. Our results serve as an encouraging proof-of-principle, but prospective clinical studies are needed to establish quantitative thresholds and evaluate performance of analysis of fragment ends for early detection, and for monitoring treatment response in patients with cancer.

## Data Availability

Plasma sequencing data generated for this study will be deposited in dbGaP once the manuscript is accepted for publication.

## Acknowledgements

We would like to thank Bethine Moore, Danielle Metz, and Stephanie Buchholtz at TGen, and the volunteers and patients who participated in this study.

## Funding

Supported by funding from the Ben and Catherine Ivy Foundation to MM, JMT and SC, from the National Cancer Institute (NCI) of the National Institutes of Health (NIH) under award number 1U01CA243078-01A1 to MM and 1R01CA223481-01 to MM, and by a Stand Up To Cancer (SU2C) – Melanoma Research Alliance Melanoma Dream Team Translational Cancer Research Grant (#SU2C-AACR-DT0612) to JMT and PML. Stand Up To Cancer is a program of the Entertainment Industry Foundation administered by the American Association for Cancer Research (AACR).

## Author Contributions

KKB, BRM and MM conceptualized and designed the study. KKB, BRM, HM, and MM developed methods. SB, JBE, BE, TKM, AS, NLT, MDP, MJB, MEB, BAP, PML, AB and JMT designed and conducted prospective clinical studies. MDS, TCC, MF, PFF and SC generated data. KKB, BRM and HM analyzed sequencing data. KKB, BRM, and MM interpreted data. KKB, BRM and MM wrote the paper with assistance from MDS, TCC, and JMT. All authors approved the final manuscript.

## Competing Interests

BRM, HM and MM are inventors on patent applications covering technologies described here including patent application number PCT/US20/41469, titled “Methods of detecting disease and treatment response in cfDNA”. MM consults for AstraZeneca and Bristol Myers Squibb, and serves on the scientific advisory board of PetDx. BRM, TCC and MM have licensing relationships with Exact Sciences. All other authors declare that they have no competing interests.

## Data and materials availability

Plasma sequencing data generated for this study will be deposited in dbGaP once the manuscript is accepted for publication. All other data associated with this study are present in the paper or the Supplementary Materials.

## Notes

### Author Declarations

Healthy volunteers were enrolled at the Translational Genomics Research Institute in Phoenix, AZ and blood samples were collected under protocol numbers 20142638, and 20181812, approved by Western Institutional Review Board (IRB). Blood and tissue samples from patients with melanoma were collected at Mayo Clinic in Scottsdale, AZ under protocol number 16-001453 and within a multi-center clinical trial (NCT02094872) under protocol number 20140190 approved by Western IRB. Blood samples from patients with breast cancer were collected at Mayo Clinic in Scottsdale, AZ under protocol number 14-006021, from patients with glioblastoma within a clinical trial (NCT02060890) at University of California in San Francisco, CA under protocol number 20141201 approved by Western IRB, and from patients with cholangiocarcinoma at Mayo Clinic in Scottsdale, AZ under protocol number 12-004713.

