## Supplementary Materials for "Analysis of fragment ends in plasma DNA from patients with cancer"

**This PDF file includes:**

Materials and Methods
Figs. S1 to S9
Tables S1 to S3

### Materials and Methods

#### Patients and samples

Healthy volunteers were enrolled at the Translational Genomics Research Institute in Phoenix, AZ and blood samples were collected under protocol numbers 20142638, and 20181812, approved by Western Institutional Review Board (IRB). Blood and tissue samples from patients with melanoma were collected at Mayo Clinic in Scottsdale, AZ under protocol number 16-001453 and within a multi-center clinical trial (NCT02094872) under protocol number 20140190 approved by Western IRB(*19*). Blood samples from patients with breast cancer were collected at Mayo Clinic in Scottsdale, AZ under protocol number 14-006021, from patients with glioblastoma within a clinical trial (NCT02060890) at University of California in San Francisco, CA under protocol number 20141201 approved by Western IRB(*20*), and from patients with cholangiocarcinoma at Mayo Clinic in Scottsdale, AZ under protocol number 12-004713. All patients provided informed consent. For a subset of patients with cancer, multiple blood samples were collected including at presentation and during treatment.

#### Sample processing, DNA extraction and sequencing

Blood samples were collected in EDTA BD Vacutainer tubes. Plasma was separated within 3 hours of venipuncture by centrifugation at 820*g* for 10 minutes, followed by a second centrifugation at 16000*g* for 10 minutes. One milliliter aliquots of plasma were stored at -80°C until DNA extraction. DNA was extracted using either MagMAX Cell-Free DNA Isolation Kit (ThermoFisher) or QIAamp Circulating Nucleic Acid Kit (Qiagen) from 1 ml to 4 ml plasma. Cell-free DNA was quantified prior to library preparation using Qubit dsDNA HS assay (ThermoFisher), Cell-free DNA ScreenTape on the TapeStation 4200 (Agilent), or using an in-house digital PCR assay(*21*). Whole genome sequencing libraries were prepared from plasma DNA using ThruPLEX Plasma-Seq or Tag-seq (Takara). Libraries were sequenced on HiSeq 4000, NextSeq 550, or NovaSeq 6000 (Illumina) to generate 75 bp to 150 bp paired-end reads.

#### Sequencing data analysis

Sequencing data was converted to fastq files using bcl2fastq v2.20.0.422. Sequencing reads were trimmed using fastp v0.20.0(*22*). Trimmed reads were aligned to human genome build hs37d5 (hg19) using bwa-mem v0.7.16a(*23*) and converted to bam files using samtools 1.9-92-gcb6b3b5(*24*). Tumor fraction was inferred using copy number analysis of plasma DNA using ichorCNA v0.3.2, together with hmmcopy for patients with melanoma and cholangiocarcinoma(*25, 26*). Reported limit of detection using ichorCNA is 3% tumor fraction. Any samples non-detectable using ichorCNA were incorporated as zeros in correlation analyses.

#### External data

Fragment end positions, and clinical annotation for patients with cancer and healthy individuals from three published studies(*25, 27, 28*) were obtained from FinaleDB(*29*). These data were processed similarly to fragment end positions identified from patients in this study.

#### Analysis of fragment ends

To analyze genomic positioning of fragment ends, a map of recurrently protected regions was inferred from 17 heathy individuals (sequenced to ~30x coverage each), using a peak-calling method based on window-protection scores(*30*). Using this map, cell-free fragments were identified as aberrant if one or both of ends were located within a protected region. Non-aberrant fragments were identified as those that span the length of a protected region. Using the counts of these two types of fragments, fraction of aberrant fragments (FAF) was calculated as the ratio of aberrant fragments to the total number of aberrant and non-aberrant fragments.

To analyze average nucleotide frequencies at fragment ends, positions from 10 bp upstream to 10 bp downstream of each fragment end were considered. For each plasma sample, average frequency across all fragments was calculated for each combination of position and base, using the sequence represented in the hg19 reference genome. Mono-nucleotide frequency was calculated at each position using samtools(*24*), BEDTools v2.29.0(*31*) and homerTools v4.11(*32*). Each sample was represented by a vector of 168 length (2 fragment ends x 4 bases x 21 positions).

For building the classification model, we used the nucleotide frequency vector and FAF for each sample. Samples were stratified by cancer type (single stratification for healthy) and split into 80% train and 20% test data. Such stratified splits ensured that train and test data share similar representation of different types of data variations, leading to improved generalization on test data(*33*). A random forest classifier (using 100 decision trees) was trained and evaluated over 100 runs and using 1000 activation thresholds uniformly distributed between 0 and 1. This binary classifier was trained using a label of 0 for healthy samples and 1 for samples from patients with cancer. The data used for building this model was limited to one sample per patient (the earliest time point available for each), to avoid potential signal leakage between train and test data.

Down-sampling analysis

To evaluate whether analysis of fragment ends was robust at lower read depths, the original datasets were subsampled using samtools(*24*). To calculate coefficient of variation for FAF, subsampling analysis was performed using earliest available plasma samples from 35 patients with melanoma. The full dataset was randomly subsampled 10 times with a maximum of 1 to 10 million fragments. With FAF computed for each random sample, coefficient of variation was calculated per observation for a given number of reads. To assess how our robust classification model was at lower read depths, a random subsample of 1 to 10 million reads was obtained from each sample included in the model and including in training and evaluation.

Comparison of FAF across copy number aberrations

To compare differences in FAF between genomics regions affected copy number aberrations, 27 plasma samples from patients with melanoma with tumor fractions of at least 20% were selected. FAF was calculated in non-overlapping 500 kb windows across the genome in each sample, along with 24 healthy control samples. For each plasma sample, we identified all windows that completely overlapped with copy number segments having less than, equal to, or greater than 2 copies. For each window, we calculated the z-score of the patient sample versus healthy controls by subtracting the mean FAF value of the bin in the healthy samples from the patient sample and dividing by the standard deviation of the healthy sample FAF values.

Comparison of FAF for mutated and non-mutated fragments

To compare FAF between mutated and non-mutated fragments, tumor and germline exome sequencing data from two patients with metastatic melanoma were analyzed, as described in an earlier study(*19*). Deep whole genome sequencing of the corresponding plasma samples was performed. Genomic loci where mutations were identified in the tumor DNA were interrogated in corresponding plasma WGS data. FAF was calculated for mutated and non-mutated fragments, in aggregate for all mutations.

Targeted digital sequencing of plasma DNA

Tumor fraction in plasma samples from patients with glioblastoma was measured using targeted digital sequencing as described earlier(*34*). Briefly, patient-specific somatic mutations were selected by analyzing exome sequencing data from tumor biopsies and germline DNA. Clonal mutations were identified, adjusting for copy number aberrations in the tumor genome and overall tumor purity. Target-specific multiplexed primers were designed and evaluated for in vitro performance using control DNA samples. TARDIS sequencing libraries were prepared and sequenced on an Illumina NovaSeq S4 flowcell. Sequencing data were analyzed to evaluate targeted genomic loci and determine confidence in ctDNA detection in each sample. ctDNA fraction was calculated as the mean of all measured variant allele fractions.

Statistical analysis

Statistical analyses were performed using Julia and Python. Significance values of differences between two FAF distributions were evaluated using the t test. Statistical significance between distribution of FAF in copy number loss, neutral, or gain regions was calculated using the Mann-Whitney U test. To compute the statistical significance of correlation, the correlation values were first converted to a t statistic and then converted to a P value based on population size. Comparison of FAF between mutated and non-mutated DNA fragments within a plasma sample was performed using the two-proportions Z test. All P values reported are two-sided. P values smaller than 0.05 were considered statistically significant.

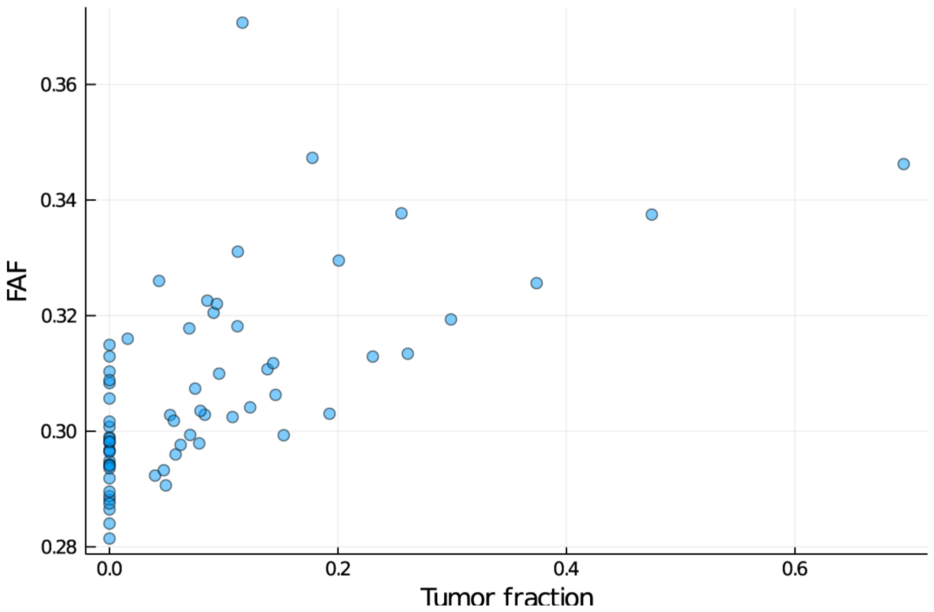


**Fig. S1.**

Comparison of tumor fraction and FAF in plasma samples from patients with cholangiocarcinoma. Tumor fraction and FAF were correlated with Pearson's r of 0.71 ($P = 2.2 \times 10^{-8}$). On the x-axis, plasma samples with tumor fraction below the limit of detection using ichorCNA are indicated as zero.

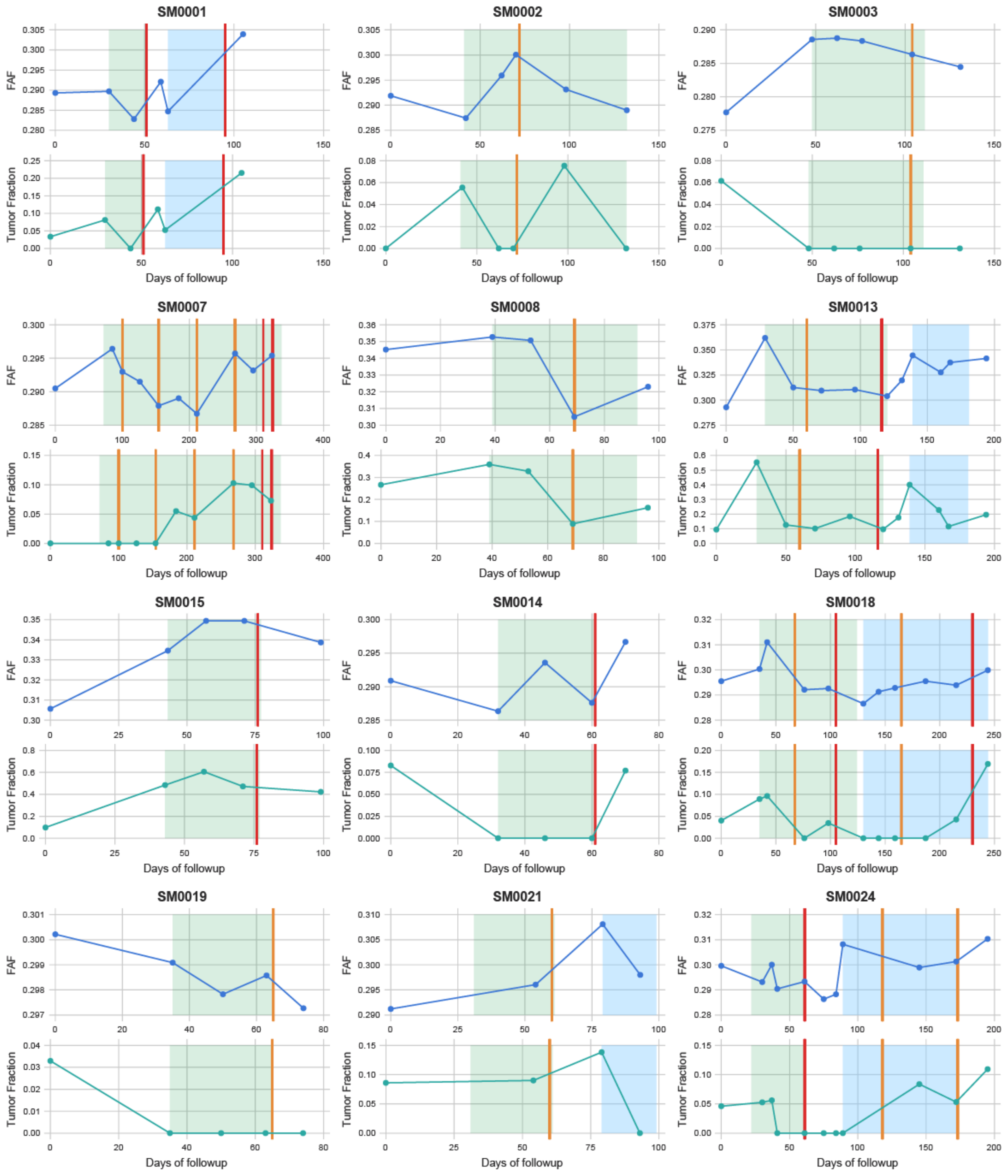
SM0001
SM0002
SM0003
SM0007
SM0008
SM0013
SM0015
SM0014
SM0018
SM0019
SM0021
SM0024
FAF
Tumor Fraction
Days of followup

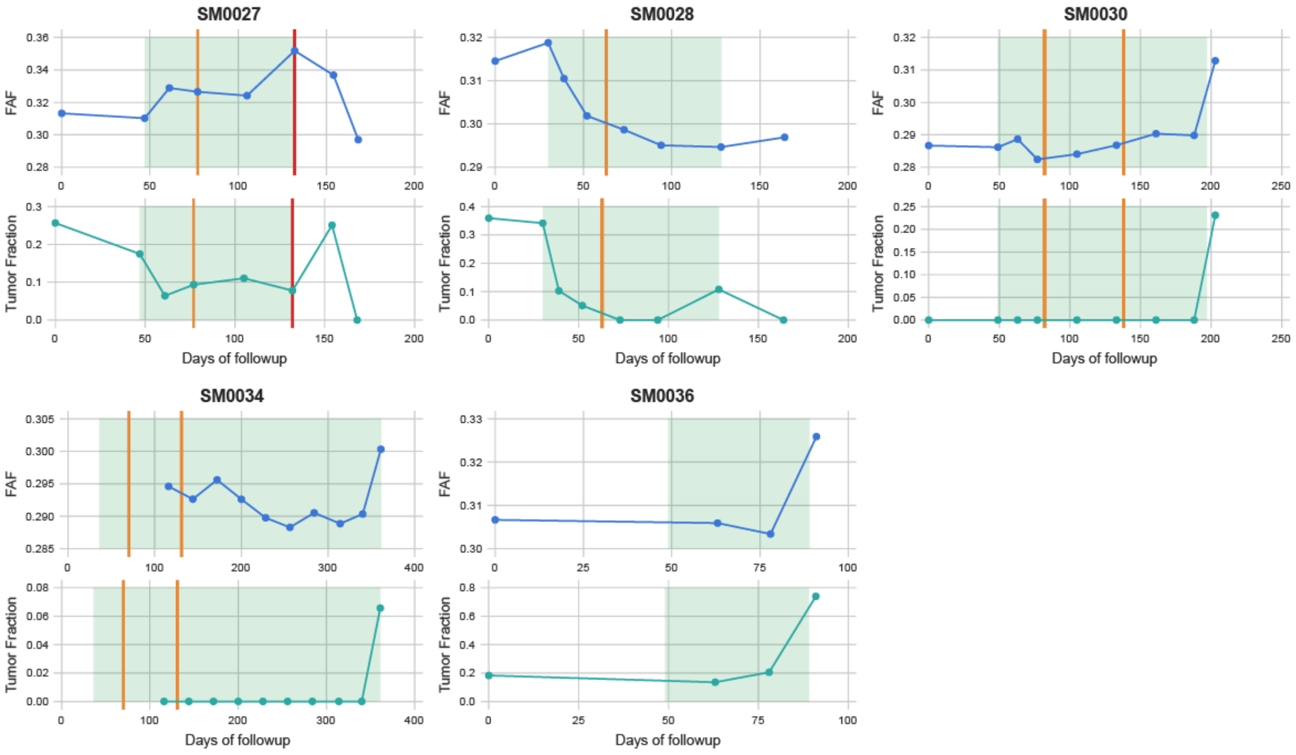


**Fig. S2.**

Comparison of longitudinal changes in tumor fraction and FAF in serial plasma samples from patients with metastatic melanoma, treated on a targeted therapy trial(*19*). 17 patients from whom at least 4 plasma samples were analyzed and at least one of them had circulating tumor DNA detectable by ichorCNA are included in this figure. For each patient, the top panel shows longitudinal changes in FAF and the bottom panel shows tumor fraction measured using ichorCNA. Days of follow-up are reported since the earliest available blood sample. Shaded areas indicate systemic therapy during the trial. When available, imaging results measured using RECIST are indicated with orange vertical lines for Stable Disease and with red vertical lines for Progressive Disease.

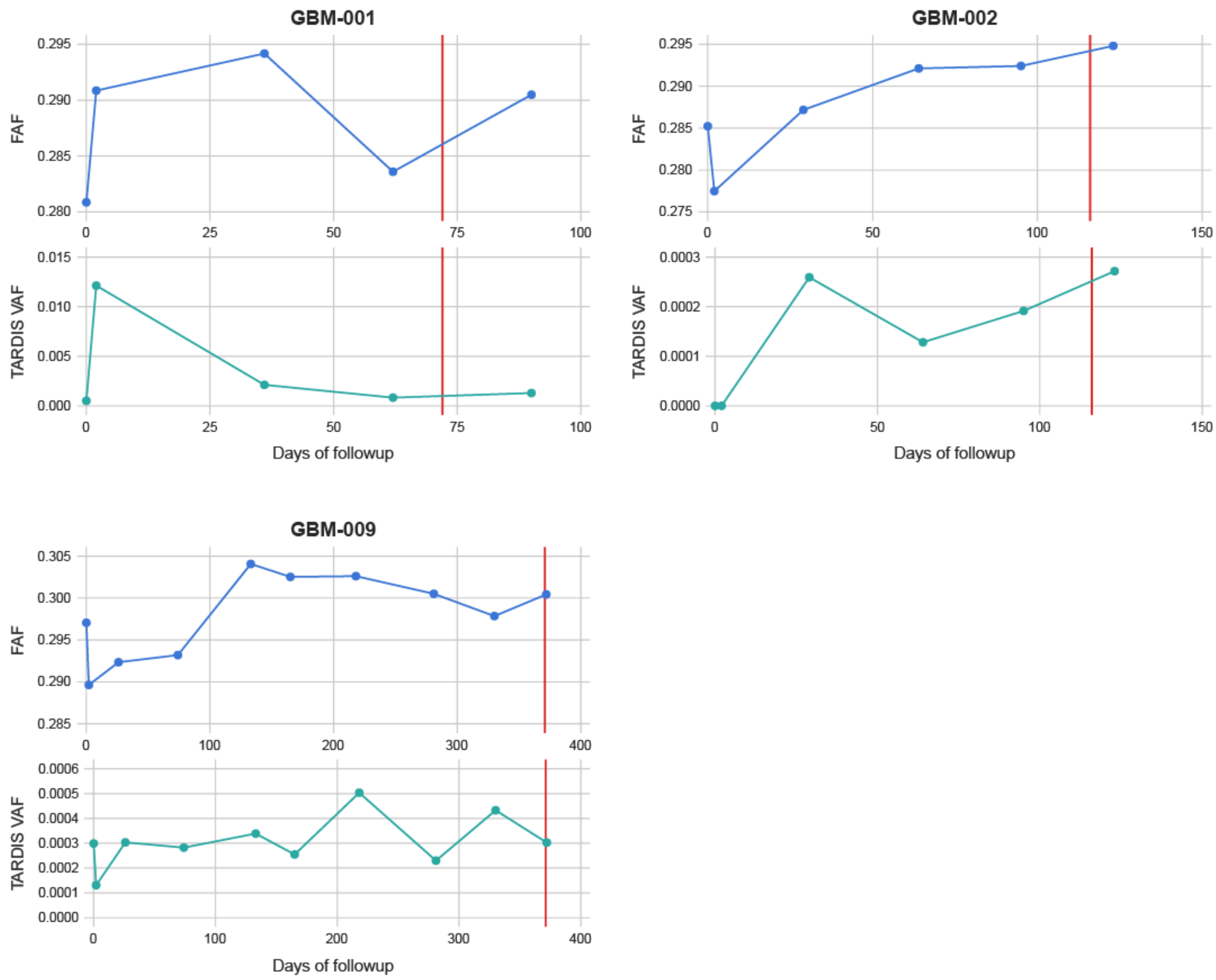


**Fig. S3.**
Comparison of longitudinal changes in tumor fraction and FAF in serial plasma samples from patients with glioblastoma, treated on a genomics-enabled therapy trial(*20*). 3 patients from whom at least 4 plasma samples were analyzed are included in this figure. For each patient, the top panel shows longitudinal changes in FAF and the bottom panel shows tumor fraction measured using TARDIS, an assay of patient-specific mutations guided by the patient's own tumor biopsy(*34*). Days of follow-up are reported since the earliest available blood sample, which was collected prior to surgical resection of the tumor. Subsequent samples were collected after surgical resection and during therapy. Vertical red line indicates clinical disease progression.

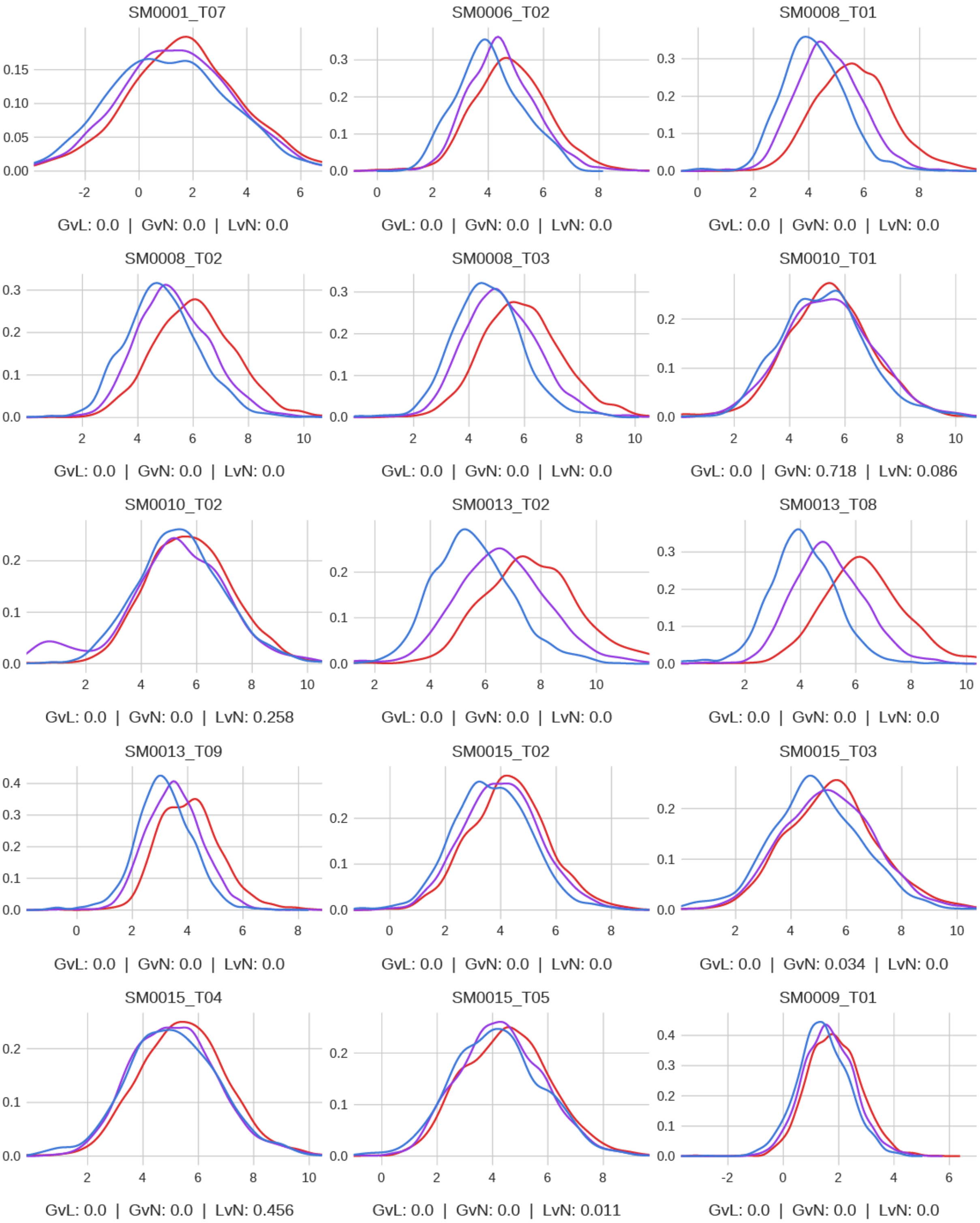
SM0001_T07
GvL: 0.0 | GvN: 0.0 | LvN: 0.0
SM0006_T02
GvL: 0.0 | GvN: 0.0 | LvN: 0.0
SM0008_T01
GvL: 0.0 | GvN: 0.0 | LvN: 0.0
SM0008_T02
GvL: 0.0 | GvN: 0.0 | LvN: 0.0
SM0008_T03
GvL: 0.0 | GvN: 0.0 | LvN: 0.0
SM0010_T01
GvL: 0.0 | GvN: 0.718 | LvN: 0.086
SM0010_T02
GvL: 0.0 | GvN: 0.0 | LvN: 0.258
SM0013_T02
GvL: 0.0 | GvN: 0.0 | LvN: 0.0
SM0013_T08
GvL: 0.0 | GvN: 0.0 | LvN: 0.0
SM0013_T09
GvL: 0.0 | GvN: 0.0 | LvN: 0.0
SM0015_T02
GvL: 0.0 | GvN: 0.0 | LvN: 0.0
SM0015_T03
GvL: 0.0 | GvN: 0.034 | LvN: 0.0
SM0015_T04
GvL: 0.0 | GvN: 0.0 | LvN: 0.456
SM0015_T05
GvL: 0.0 | GvN: 0.0 | LvN: 0.011
SM0009_T01
GvL: 0.0 | GvN: 0.0 | LvN: 0.0

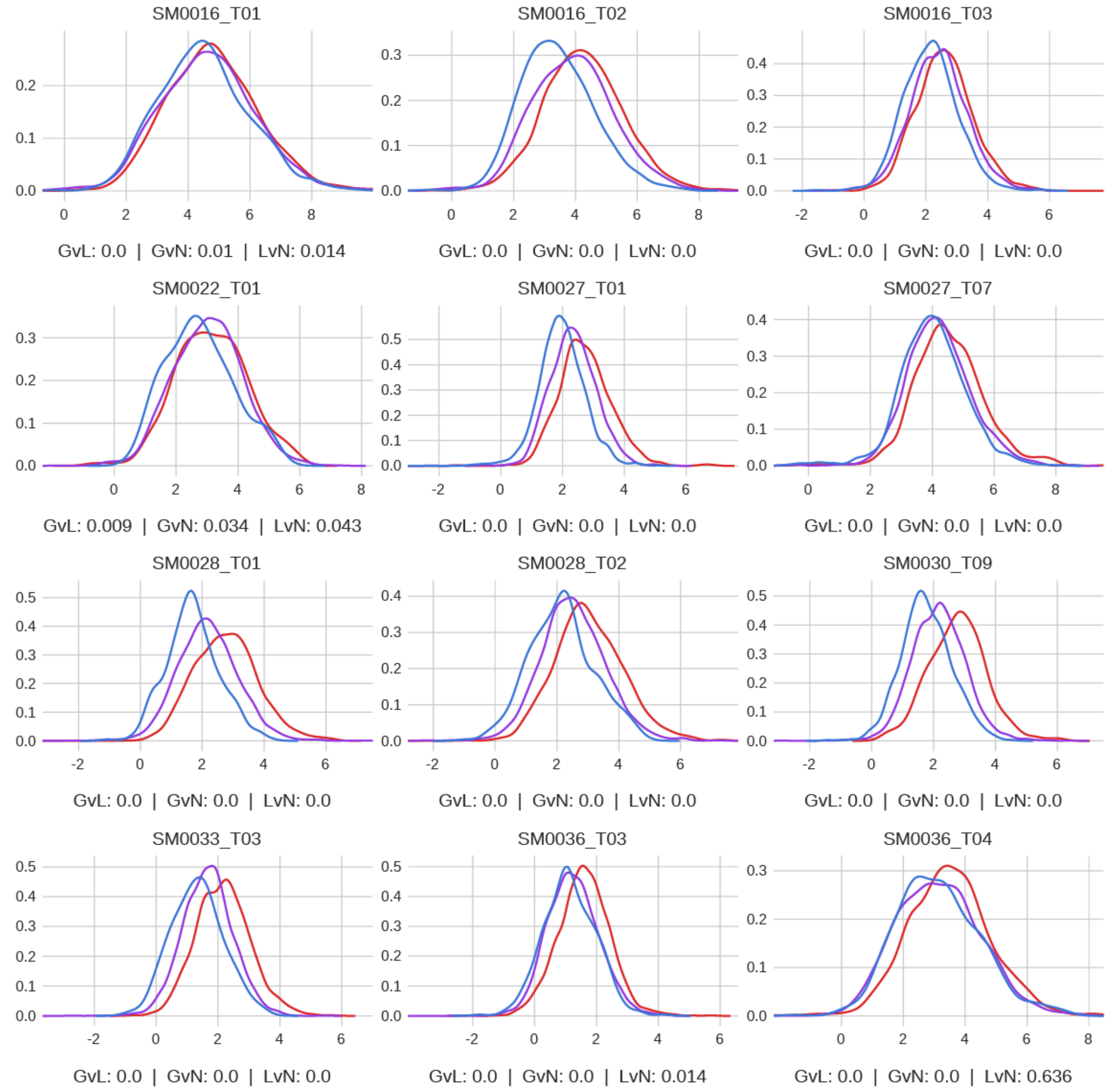


**Fig. S4.**

Comparison of FAF between copy number gain, neutral and loss regions in patients with metastatic melanoma. Density plots for normalized FAF are presented for copy number loss (blue), neutral (purple) and gain regions (red) for 27 plasma samples with at least 20% tumor fraction measured using ichorCNA. Under each plot, p values for comparison of these distributions are presented. GvL: gain regions vs. loss regions. GvN: gain regions vs. neutral regions. LvN: loss regions vs. neutral regions. All 27 samples showed significantly higher FAF in gain regions compared to neutral regions, in gain regions compared to loss regions, or both ($P < 0.05$).

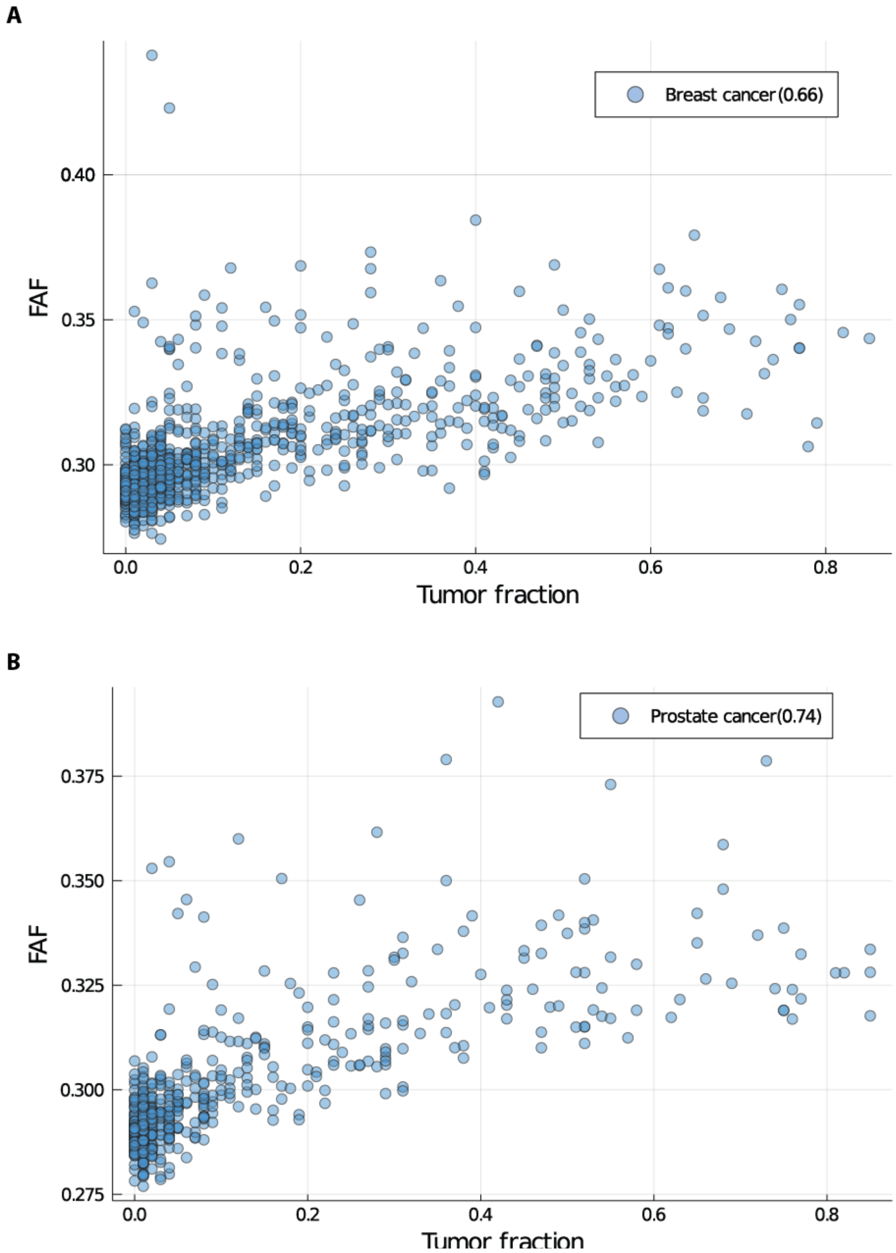


**Fig. S5.**

Comparison of tumor fraction and FAF in plasma samples from patients with metastatic breast and prostate cancer. Whole genome sequencing data from Adalsteinsson et al. was analyzed for this figure(*25*). Tumor fraction and FAF were correlated with Pearson's r of 0.66 ($P = 1.9 \times 10^{-119}$) in plasma samples from patients with metastatic breast cancer (A) and with Pearson's r of 0.74 ($p = 6.8 \times 10^{-98}$) in plasma samples from patients with metastatic prostate cancer (B). On the x-axis, plasma samples with tumor fraction below the limit of detection using ichorCNA are indicated as zero.

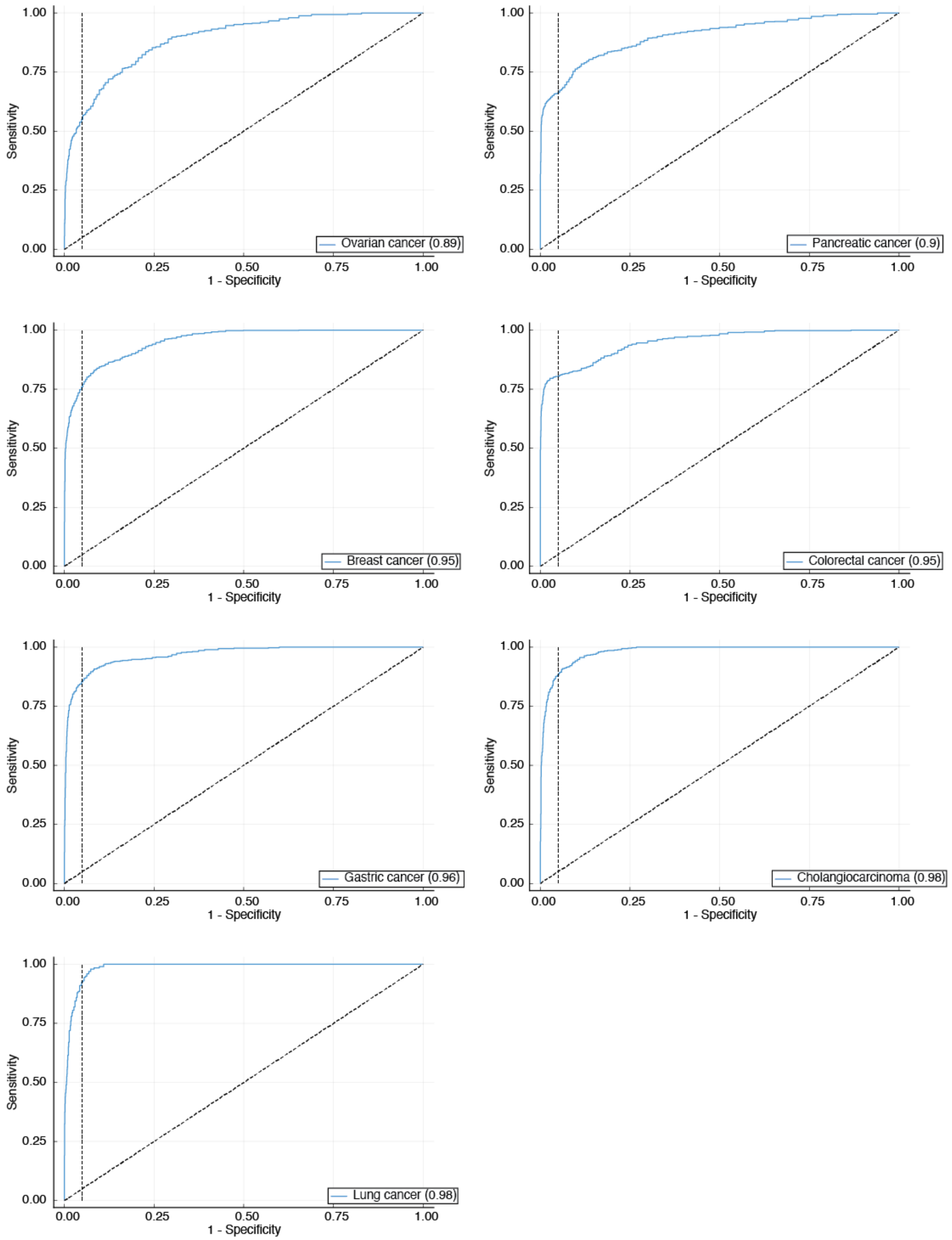
Ovarian cancer (0.89)
Pancreatic cancer (0.9)
Breast cancer (0.95)
Colorectal cancer (0.95)
Gastric cancer (0.96)
Cholangiocarcinoma (0.98)
Lung cancer (0.98)
Sensitivity
1 - Specificity
0.00
0.25
0.50
0.75
1.00

**Fig. S6.**

ROC curves for cancer detection by cancer type. Whole genome sequencing data from Cristiano et al. was used to evaluate performance of analysis of fragment ends(*27*). Each panel shows classifier performance in a cancer subtype. Numbers with brackets are areas under the ROC curves.

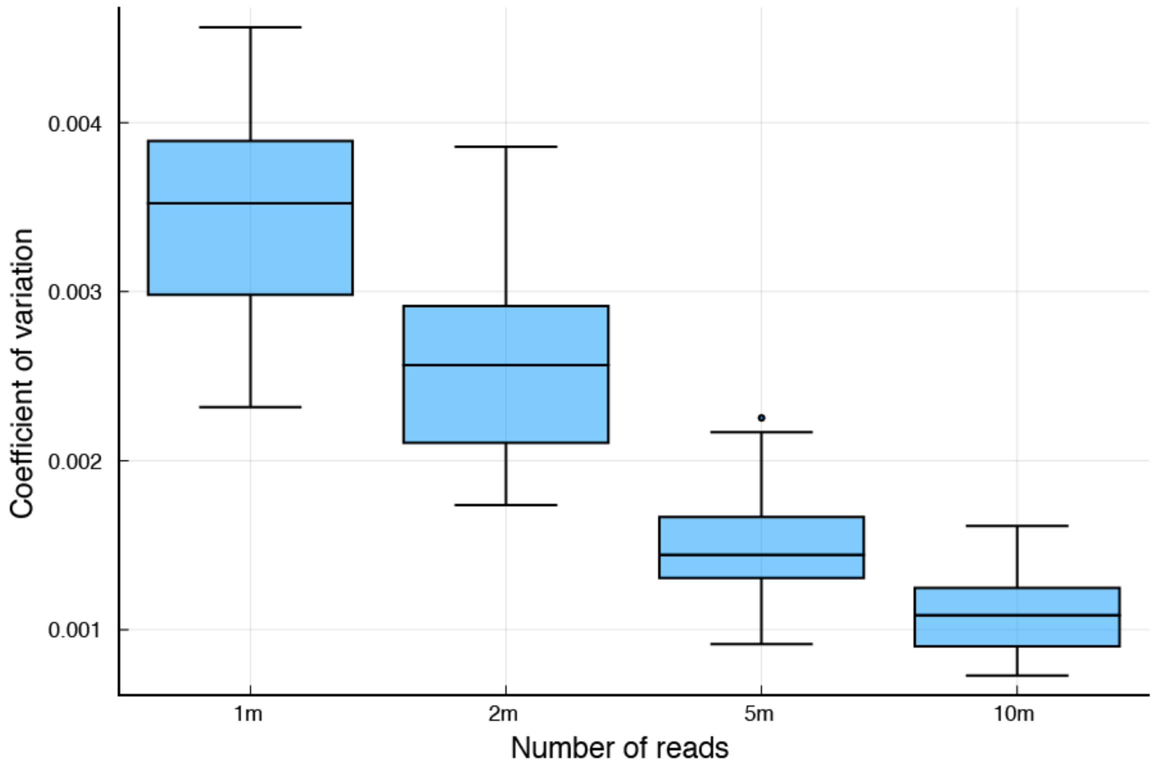


**Fig. S7.**
Co-efficient of variation (CV) for FAF in down-sampled data sets. To calculate CV, multiple independent datasets with decreasing number of DNA fragments were generated and FAF was calculated from these replicates. CVs remained less 1% even for as low as 1 million reads per sample.

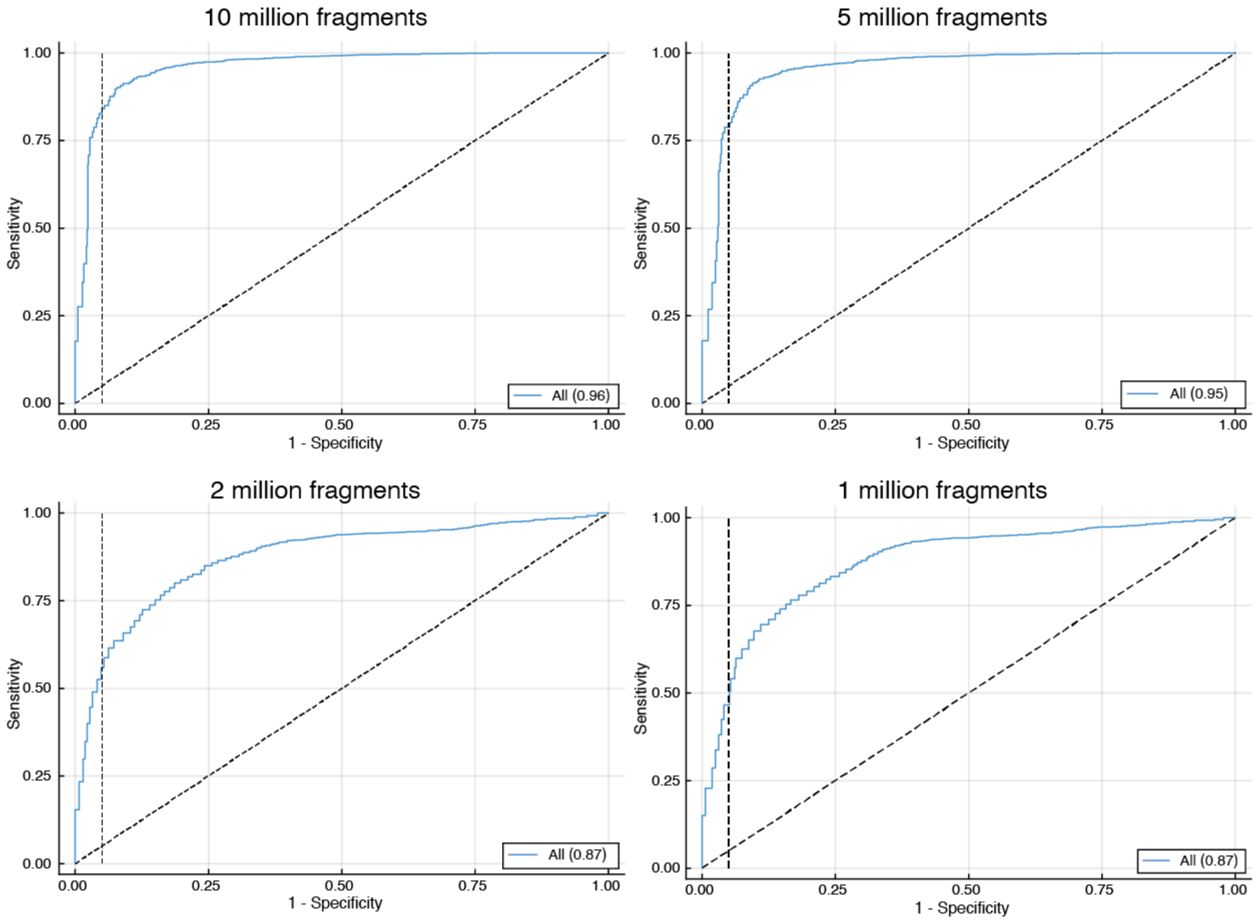


**Fig. S8.**

Classifier performance with down-sampling in our multi-cancer cohort. Down-sampling was performed to limit maximum number of analyzed fragments, as indicated on each panel. Overall classifier performance for cancer detection is shown. Numbers in brackets are area under the ROC curve. Vertical dashed black line indicates 95% specificity.

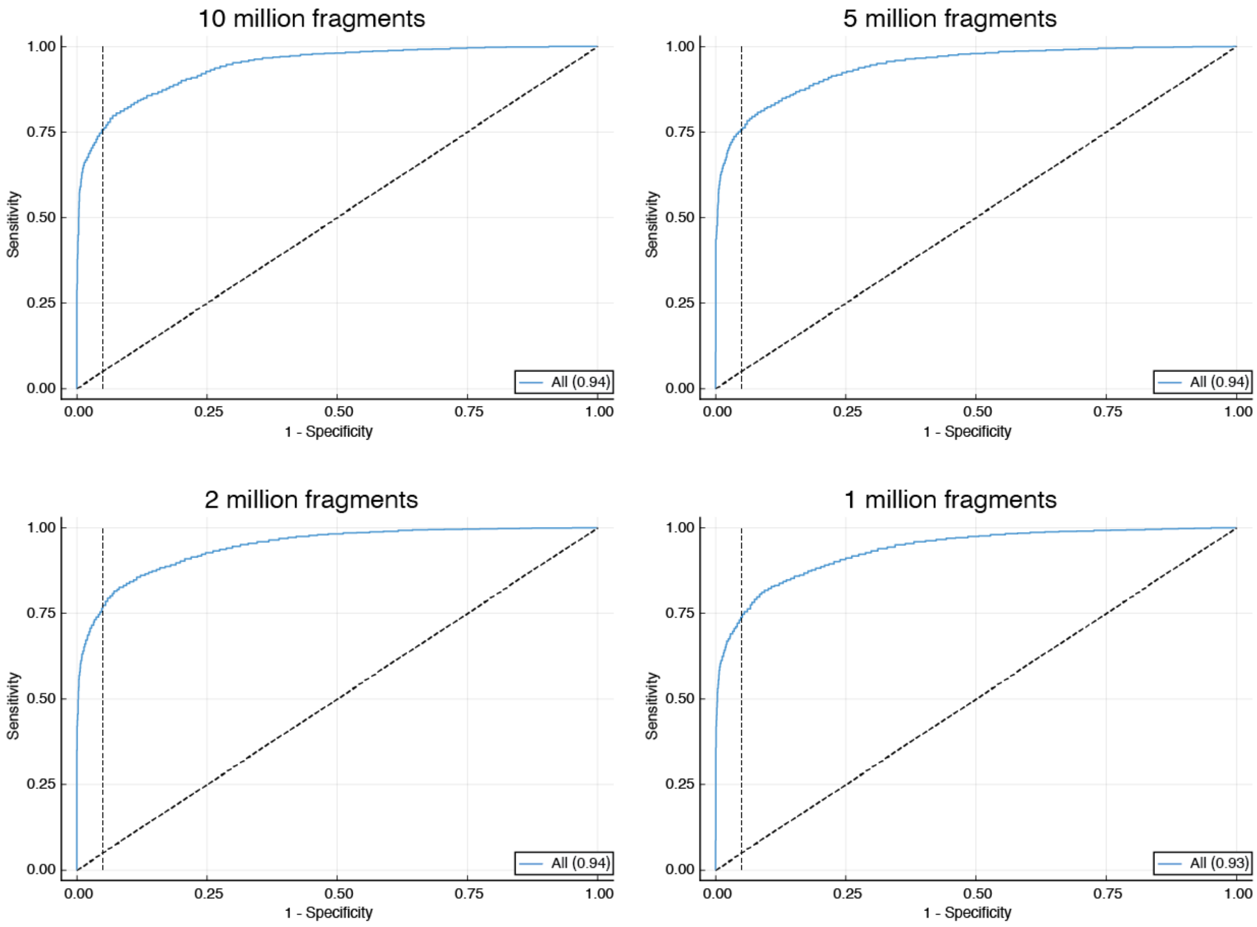


**Fig. S9.**

Classifier performance with down-sampling in Cristiano et al.'s published cohort(*27*). Down-sampling was performed to limit maximum number of analyzed fragments, as indicated on each panel. Overall classifier performance for cancer detection is shown. Numbers in brackets are area under the ROC curve. Vertical dashed black line indicates 95% specificity.

| Study | Diagnosis | Number of Samples | Mean FAF | Standard Deviation of FAF | P value | Cohen's d |
|---|---|---|---|---|---|---|
| This study | Healthy Individuals | 40 | 0.287 | 0.004 | - | - |
| | Breast Cancer | 47 | 0.299 | 0.012 | $4.5x10^{-8}$ | 1.3 |
| | Cholangiocarcinoma | 46 | 0.307 | 0.016 | $9.4x10^{-11}$ | 1.6 |
| | Glioblastoma | 45 | 0.301 | 0.013 | $9.3x10^{-9}$ | 1.4 |
| | Melanoma | 261 | 0.299 | 0.016 | $3.3x10^{-6}$ | 0.8 |
| Cristiano et al.(*27*) | Healthy Individuals | 262 | 0.285 | 0.005 | - | - |
| | Breast Cancer | 54 | 0.290 | 0.008 | $6.9x10^{-9}$ | 1.1 |
| | Cholangiocarcinoma | 25 | 0.293 | 0.006 | $4.7x10^{-11}$ | 1.9 |
| | Colorectal Cancer | 27 | 0.298 | 0.014 | $4.7x10^{-21}$ | 2.4 |
| | Gastric Cancer | 27 | 0.288 | 0.012 | $8.2x10^{-3}$ | 0.7 |
| | Lung Cancer | 79 | 0.293 | 0.009 | $1.5x10^{-21}$ | 3.0 |
| | Ovarian Cancer | 28 | 0.292 | 0.009 | $1.4x10^{-10}$ | 1.7 |
| | Pancreatic Cancer | 35 | 0.290 | 0.007 | $8.0x10^{-8}$ | 1.4 |
| Jiang et al.(*28*) | Healthy Individuals | 32 | 0.281 | 0.003 | - | - |
| | Liver Cirrhosis | 36 | 0.281 | 0.004 | 0.65 | - |
| | Hepatitis B | 67 | 0.280 | 0.004 | 0.52 | - |
| | Hepatocellular Carcinoma | 90 | 0.288 | 0.010 | $1.5x10^{-4}$ | 0.8 |
| Adalsteinsson et al.(*25*) | Breast Cancer | 950 | 0.304 | 0.019 | $1.5x10^{-8}$ | 0.9 |
| | Prostate Cancer | 558 | 0.301 | 0.017 | $9.7x10^{-7}$ | 0.8 |

**Table S1.**

Comparison of FAF between analyzed samples and cohorts. For each study, groups of patients were compared with data from the study's corresponding healthy individual samples. For Adalsteinsson et al., no healthy individual sample data was available and patient groups were compared with healthy individuals in our study. Two-tailed p values are reported from Student's t-test. No significant elevation in FAF was observed for patients with liver cirrhosis or hepatitis B.

| Diagnosis | Patient ID | Sample ID | Number of non-mutated fragments | FAF in non-mutated fragments | Number of mutated fragments | FAF in mutated fragments | P value |
|---|---|---|---|---|---|---|---|
| Melanoma | SM0008 | SM0008_T02 | 248405 | 0.334 | 79737 | 0.364 | $<2.2x10^{-16}$ |
| Melanoma | SM0022 | SM0022_T01 | 297031 | 0.307 | 76809 | 0.328 | $1.5x10^{-11}$ |

**Table S2.**
Comparison of aberrant positioning between mutated and non-mutated fragments. Two-tailed p-values are reported from two proportions Z test.

| | | | Nucleotide frequencies at fragment ends | |
|---|---|---|---|---|
| | | | Dimension 1 | Dimension 2 |
| Correlation with tumor fraction | This study | CCA | -0.169 | 0.551 |
| | | Melanoma | 0.085 | 0.551 |
| | Adalsteinsson et al. | Breast Cancer | 0.002 | -0.571 |
| | | Prostate Cancer | 0.099 | -0.447 |
| Correlation with FAF | This study | CCA | 0.108 | 0.735 |
| | | Melanoma | 0.183 | 0.700 |
| | Adalsteinsson et al. | Breast Cancer | 0.004 | -0.712 |
| | | Prostate Cancer | 0.106 | -0.476 |

**Table S3.**
Correlation of nucleotide frequencies at fragment ends with tumor fraction and FAF in plasma DNA. Correlation between dimension 2 of nucleotide frequencies at fragment ends with tumor fraction and with FAF were all statistically significant ($P < 0.05$).

19. P. M. LoRusso *et al.*, Identifying treatment options for BRAFV600 wild-type metastatic melanoma: A SU2C/MRA genomics-enabled clinical trial. *PLoS One* **16**, e0248097 (2021).
20. S. A. Byron *et al.*, Prospective Feasibility Trial for Genomics-Informed Treatment in Recurrent and Progressive Glioblastoma. *Clin Cancer Res* **24**, 295-305 (2018).
21. H. Markus *et al.*, Evaluation of pre-analytical factors affecting plasma DNA analysis. *Sci Rep* **8**, 7375 (2018).
22. S. Chen, Y. Zhou, Y. Chen, J. Gu, fastp: an ultra-fast all-in-one FASTQ preprocessor. *Bioinformatics* **34**, i884-i890 (2018).
23. H. Li, Aligning sequence reads, clone sequences and assembly contigs with BWA-MEM. *arXiv preprint arXiv:1303.3997*, (2013).
24. H. Li *et al.*, The Sequence Alignment/Map format and SAMtools. *Bioinformatics* **25**, 2078-2079 (2009).
25. V. A. Adalsteinsson *et al.*, Scalable whole-exome sequencing of cell-free DNA reveals high concordance with metastatic tumors. *Nat Commun* **8**, 1324 (2017).
26. G. Ha *et al.*, Integrative analysis of genome-wide loss of heterozygosity and monoallelic expression at nucleotide resolution reveals disrupted pathways in triple-negative breast cancer. *Genome Res* **22**, 1995-2007 (2012).
27. S. Cristiano *et al.*, Genome-wide cell-free DNA fragmentation in patients with cancer. *Nature* **570**, 385-389 (2019).
28. P. Jiang *et al.*, Lengthening and shortening of plasma DNA in hepatocellular carcinoma patients. *Proc Natl Acad Sci U S A* **112**, E1317-1325 (2015).
29. H. Zheng, M. S. Zhu, Y. Liu, FinaleDB: a browser and database of cell-free DNA fragmentation patterns. *Bioinformatics*, (2020).
30. M. W. Snyder, M. Kircher, A. J. Hill, R. M. Daza, J. Shendure, Cell-free DNA Comprises an In Vivo Nucleosome Footprint that Informs Its Tissues-Of-Origin. *Cell* **164**, 57-68 (2016).
31. A. R. Quinlan, I. M. Hall, BEDTools: a flexible suite of utilities for comparing genomic features. *Bioinformatics* **26**, 841-842 (2010).
32. S. Heinz *et al.*, Simple combinations of lineage-determining transcription factors prime cis-regulatory elements required for macrophage and B cell identities. *Mol Cell* **38**, 576-589 (2010).
33. N. Wan *et al.*, Machine learning enables detection of early-stage colorectal cancer by whole-genome sequencing of plasma cell-free DNA. *BMC Cancer* **19**, 832 (2019).
34. B. R. McDonald *et al.*, Personalized circulating tumor DNA analysis to detect residual disease after neoadjuvant therapy in breast cancer. *Sci Transl Med* **11**, (2019).
